# Clinical evaluation of the Abbott Alinity SARS-CoV-2 spike-specific quantitative IgG and IgM assays in infected, recovered, and vaccinated groups

**DOI:** 10.1101/2021.02.17.21251940

**Authors:** Madhusudhanan Narasimhan, Lenin Mahimainathan, Ellen Araj, Andrew E. Clark, John Markantonis, Allen Green, Jing Xu, Jeffrey A. SoRelle, Charles Alexis, Kimberly Fankhauser, Hiren Parikh, Kathleen Wilkinson, Annika Reczek, Noa Kopplin, Sruthi Yekkaluri, Jyoti Balani, Abey Thomas, Amit Singal, Ravi Sarode, Alagarraju Muthukumar

**Affiliations:** Department of Pathology, University of Texas Southwestern Medical Center, Dallas, Texas; Department of Internal Medicine, University of Texas Southwestern Medical Center, Dallas, Texas

**Keywords:** COVID-19, SARS-CoV-2, IgG, IgM, Spike, Nucleocapsid, Vaccine

## Abstract

The COVID-19 pandemic continues to impose a significant burden on global health infrastructure. While identification and containment of new cases remains important, laboratories must now pivot and consider assessment of SARS-CoV-2 immunity in the setting of the recent availability of multiple COVID-19 vaccines. Here we have utilized the latest Abbott Alinity semi-quantitative IgM and quantitative IgG spike protein (SP) serology assays (IgM_SP_ and IgG_SP_) in combination with Abbott Alinity IgG nucleocapsid (NC) antibody test (IgG_NC_) to assess antibody responses in a cohort of 1236 unique participants comprised of naïve, SARS-CoV-2 infected, and vaccinated (including both naïve and recovered) individuals. The IgM_SP_ and IgG_SP_ assays were highly specific (100%) with no cross-reactivity to archived samples recovered prior to the emergence of SARS-CoV-2, including those from individuals with seasonal coronavirus infections. Clinical sensitivity was 96% after 15 days for both IgM_SP_ and IgG_SP_ assays individually. When considered together, the sensitivity was 100%. A combination of NC- and SP-specific serologic assays clearly differentiated naïve, SARS-CoV-2-infected, and vaccine-related immune responses. Vaccination resulted in a significant increase in IgG_SP_ and IgM_SP_ titers, with a major rise in IgG_SP_ following the booster (second) dose in the naïve group. In contrast, SARS-CoV-2 recovered individuals had several fold higher IgG_SP_ responses than naïve following the primary dose, with a comparatively dampened response following the booster. This work illustrates the strong clinical performance of these new serological assays and their utility in evaluating and distinguishing serological responses to infection and vaccination.

## Introduction

Severe acute respiratory syndrome coronavirus 2 (SARS-CoV-2) is a novel human pathogen that causes coronavirus disease 19 (COVID-19). The global spread of the virus began in early 2020 and continues to dramatically impact global public health infrastructure. Until recently, identification of positive cases, treatment of a patient with severe or life-threatening symptomology, and containment to prevent disease transmission have been the most widely utilized epidemiological tools used. However, the recent availability of vaccines engineered to provide protective humoral immunity represents a major step forward in the national response to SARS-CoV-2, and a pivoting point for the clinical laboratory.

Laboratory diagnostic focus must now expand to include a more thorough assessment of SARS-CoV-2 immunity due to vaccine availability. The molecular detection of SARS-CoV-2 RNA remains the gold standard for disease diagnosis. These methods are highly sensitive in the immediate period post-symptom onset, but diagnostic sensitivity diminishes by about 50% after one week of symptoms (1-3). Conversely, the sensitivity of serological assays improves following two weeks after symptom onset (2,4), making these methods helpful for establishing disease prevalence in a population.

Akin to natural infection, the process of vaccination stimulates the immune system to form memory B-cells that produce neutralizing antibodies (Ab). The two successful messenger RNA (mRNA)-based COVID-19 vaccines approved and administered in the United States contain mRNA that encodes the spike (SP) protein of SARS-CoV-2 (Pfizer-BioNTech and Moderna). Serological methods are also important for their roles in assessing immune status in vaccinated and unvaccinated individuals. However, to accurately address these questions, the selected serological tests must exhibit exceptional sensitivity and specificity with minimal cross-reactivity.

In this work, we evaluated the clinical performance of two new serological assays designed to quantitatively evaluate the presence of specific IgM (IgM_SP_) and IgG (Abbott SARS-CoV-2 IgG II; IgG_SP_) recognizing SARS-CoV-2 spike protein. Additionally, we evaluated the use of these assays in combination with an assay which detects IgG specific to the nucleocapsid protein (IgG_NC_) assay (Abbott) in SARS-CoV-2-infected, naïve/uninfected, and vaccinated groups. In addition to exhibiting excellent analytical characteristics, we found the combination of these spike-protein and nucleocapsid-specific assays as an excellent tool for the assessment of serological responses to prior infection versus vaccination.

## Materials and Methods

### Patient samples

A total of 1236 individuals (1428 specimens) were used in this study. This included 263 inpatients with suspected SARS-CoV-2 infection (413 samples), 404 unique patients for specificity and cross-reactivity studies including, 338 patients with samples obtained prior to the emergence of SARS-CoV-2, 569 individuals (611 samples) tested in outpatient setting with COVID-19 vaccination information available (Fig. 1). For the assessment of clinical sensitivity, only inpatients with suspected SARS-CoV-2 infection with PCR and documented dates of symptom onset were included. In order to accurately evaluate the serological response following vaccination, only outpatients with information concerning their vaccine status were included. This study was approved by the Institutional Review Board of the University of Texas Southwestern Medical Center.

**FIG 1.**
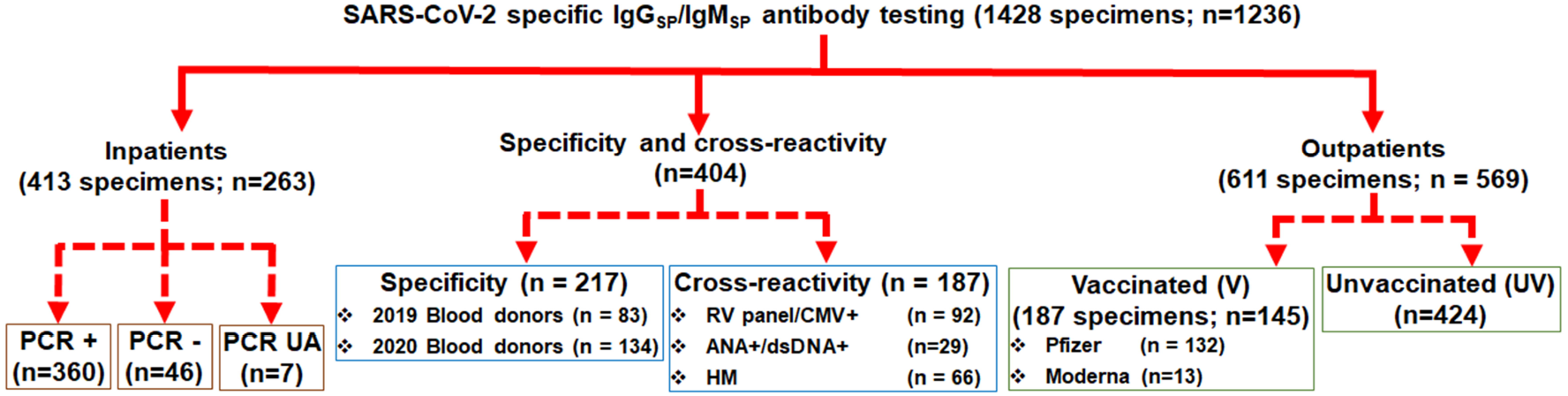
Flowchart of cases used in the study. Samples from one thousand two hundred and thirty-six unique individuals were used for SARS-CoV-2-IgGSP and IgMSP testing. This includes a total of 413 specimens from 263 unique inpatient samples that was further confirmed by RT-PCR and this included 360 PCR positive for SARS-COV-2, 46 PCR negative cases, and 7 unavailable data; 217 (collected during 2019-2020 pre-COVID-19 period in the United States) and 92 HLA lab-confirmed (collected during 2015-2019) respiratory panel positive, 29 lupus patient samples, and 66 HM samples (collected during COVID period) that were used for specificity and cross-reactivity analysis, respectively; 569 outpatient cases with data clearly available for vaccination (n=145) and unvaccinated (n=424). HLA, human leukocyte antigen; PCR, polymerase chain reaction; +, positive for SARS-CoV-2; -, negative for SARS-CoV-2; UA, unavailable; Ig, immunoglobulins; NC, nucleocapsid; SP, spike; RV, respiratory viral; CMV, cytomegalovirus; ANA+, anti-nuclear antibodies positive; dsDNA+, anti-double stranded DNA positive; HM, hematological malignancies, V, vaccinated, UV, unvaccinated.

### Analytical precision

Precision testing was performed in accordance with CLIA guidelines and in accordance with standard laboratory practices at the University of Texas Southwestern Medical Center. A typical directed 5-day precision study following CLSI EP5-A3 guidelines was conducted. Two levels of vendor provided quality controls were tested with 5 replicates in the morning and 5 replicates in the afternoon for 5 days for each test.

### Measurement range and functional sensitivity

Linearity (analytical measurement range) testing was performed as a single day study following CLSI EP6-A. Patient samples with elevated IgG_SP_ and IgM_SP_ values were chosen to prepare linearity panels per CLSI EP6-A and according to the manufacturer’s guidelines. Linearity panels consisted of 7-8 levels covering the linear range of the test, with one level within the 10^th^ percentile of the lower reportable range, one level with the 90^th^ percentile of the upper reportable range and one level around the cut-off range of the assays. A minimum of three replicates were tested for each level.

Clinical reportable range was calculated by diluting samples with elevated values that were just below the upper analytical measurement range of the assays at 1:2 and 1:4 dilutions, and comparing the results to their neat (undiluted) sample results (≤ 10% acceptability criteria). Functional sensitivity of the quantitative IgG_SP_ test (7 AU/mL) was verified by running a neat patient sample (8.3 AU/mL) and its 1:1 dilution (4.15 AU/mL, expected value) ten times each and calculating their % CV (acceptable criteria ≤ 20% imprecision) as recommended by the manufacturer.

### Carry over and sample stability studies

Carry over studies were undertaken with a positive (high sample) and negative control (low sample). The high sample was run three times followed by three runs of low sample and this was repeated three times. Ten patient samples including both positive and negative for SARS-CoV-2 were tested repeatedly over a period of 12 days for specimen stability analysis. An acceptance criterion of 10% or an index value of 0.1 for IgM_SP_ and 5 AU/ml for IgG_SP_ were utilized.

### IgM_SP_ assay

The AdviseDx SARS-CoV-2 IgM_SP_ assay has recently been granted Emergency Use Authorization by the US Food and Drug administration. IgM_SP_ testing was performed on the Abbott Alinity i platform per manufacturer instructions. The test is a chemiluminescent microparticle (CMIA) assay for semi-quantitative assessment of IgM antibodies to the spike protein of SARS-CoV-2 in human serum and plasma sample. A vendor recommended cut-off of 1.0 (index value) for reactivity/positivity of infection (FDA approval for CoV-2 IgM; 6R87; H14977R01) was applied.

### IgG_SP_ assay

SARS-CoV-2 IgG II quantitative testing (under FDA review) was performed on the Abbott Alinity i platform in accordance with manufacturer’s package insert. In this antibody CMIA test, the SARS-CoV-2 antigen coated paramagnetic microparticles bind to the IgG antibodies that attach to the virus’ spike protein in human serum and plasma sample. The resulting chemiluminescence in relative light units (RLU) following the addition of anti-human IgG-labeled in comparison with the IgG II calibrator/standard indicates the strength of response, which reflects the quantity of IgG_SP_ present. 50 AU/mL and above in this test are considered as positive. This quantitative measurement of IgG_SP_ can be helpful to evaluate an individual’s humoral response to vaccines.

### IgG_NC_ assay

The Abbott Alinity i SARS-CoV-2 anti-nucleocapsid protein IgG assay is a semi-quantitative CMIA assay that has been previously verified for routine patient testing in our institution’s clinical laboratory, and the index values of 1.4 and above is considered as positive (5) per the manufacturer’s instructions.

### RT-PCR testing

The Abbott M2000 or Abbott Alinity M RT-PCR-based molecular testing/confirmation for SARS-CoV-2 was performed using nasopharyngeal specimens collected in viral transport media as previously described (5).

### Clinical Specificity

Specificity was evaluated in the pre-COVID-19 era remnant banked plasma samples from 217 unique patients collected from blood donors September to November, 2019 and from March to April 2020.

### Cross-reactivity studies

The serum/plasma specimens for cross-reactivity studies included HLA lab-confirmed positive samples (collected from January 1, 2015 to September 30, 2019) for CMV IgG, influenza A/B, respiratory syncytial virus, and endemic human coronaviruses (n=92). Also, the samples that typically contain significant levels of antibodies such as lupus patients (n=29; collected between 2004-2007) positive for anti-nuclear antibodies (ANA) and anti-double stranded DNA, and hematological malignancies (HM) (n=66; collected between March and October 2020) were included.

### Statistical analysis

Data analysis was carried out using GraphPad Prism software (Version 9,0.1, San Diego, CA, USA). Data are presented as median with range. When experiments involved more than two groups, one-way ANOVA followed by Tukey multiple comparison post-hoc analysis was used to analyze the statistical differences. For experiments involving only two groups, as appropriate, paired or unpaired Student’s t-test was performed. A p value less than 0.05 (<0.05) was considered statistically significant.

## Results

### Analytical performance

For both the IgG_SP_ and IgM_SP_ assays, the imprecision was less than 5%. The analytical measurement range of the IgG_SP_ assay was from 4.2 −50,000 AU/mL (clinical reportable range up to 200,000 AU/mL) and for IgM_SP_ assay we observed a linearity up to 16 index value. Manufacturer recommended functional sensitivity of 7 AU/mL of the quantitative IgG_SP_ assay was confirmed. There were no carry-over issues with both the assays. The samples kept refrigerated were stable over 12 days test period.

### Clinical specificity and cross-reactivity

Clinical specificity of the assay was evaluated with serum/plasma from blood donors (SARS-CoV-2 uninfected) obtained before the emergence of SARS-CoV-2. None of these samples were positive (0/217; 100% specificity) for SARS-CoV-2 spike-specific IgG (Fig. 2A) and spike-specific IgM (Fig. 2B) that matched with the manufacturer’s claimed specificity. The median IgG_SP_ value and IgM_SP_ index value were found to be 1.92 and 0.05, respectively, well below the corresponding positive cut-off limit of 50 AU/mL for IgG_SP_ and 1.0 index value for IgM_SP_.

**FIG 2.**
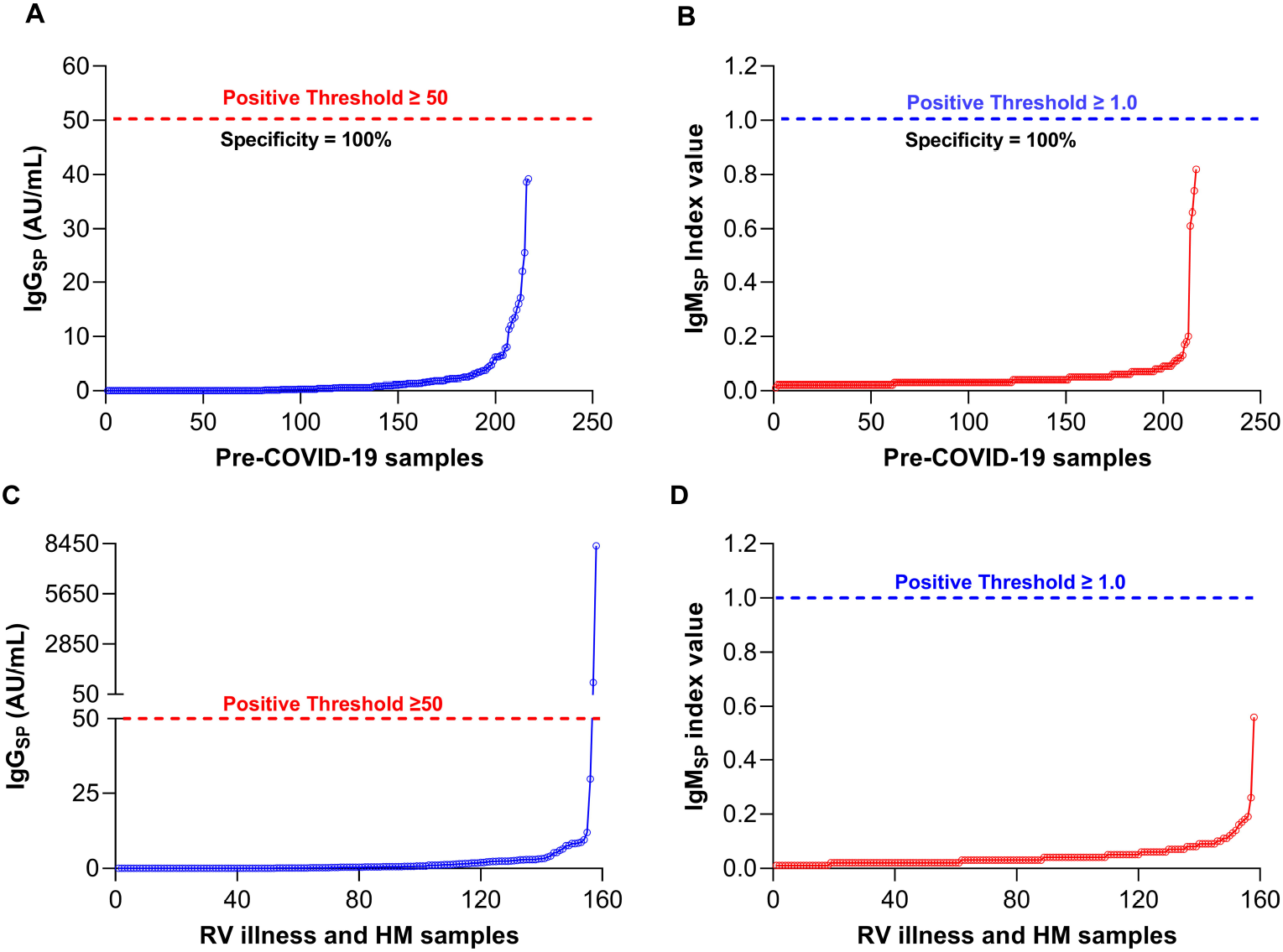
Specificity and cross-reactivity of Abbott SARS-CoV-2 IgGSP and IgMSP Assays. (A) Specificity of IgG_SP_ assay in samples from pre-COVID-19 period. (B) Specificity of IgM_SP_ assay in samples from pre-COVID-19 period; (C) Cross-reactivity of IgG_SP_ assay; (D) Cross-reactivity of IgM_SP_ analysis.

Cross-reactivity of the Abbott SARS-CoV-2 IgG_SP_ and IgM_SP_ assays was also evaluated in the setting of respiratory illness and disease conditions, including lupus and hematological malignancies (HM) (primarily multiple myeloma) samples, where antibody production is significantly elevated (Figs. 2C & 2D). 121 samples obtained prior to SARS-CoV-2 emergence did not cross-react with IgG_SP_. Interestingly, among 66 HM patient samples collected in 2020, 2 patients (∼1.1%) were found to produce a positive value for IgG_SP_ indicating some sort of cross-reactivity (Fig. 2C). One of these cases (a multiple myeloma patient) was confirmed by PCR as having recovered from COVID-19 upon chart review. Interestingly, the second positive patient was repeatedly negative for SARS-CoV-2 by PCR and strongly positive for Hepatitis viral panel serology testing. As no history of SARS-CoV-2 exposure could be established for this patient, further investigation of this IgG_SP_-specific apparent cross-reactivity is warranted. No cross-reactivity was observed in the IgM_SP_ assay (0/187 patients) (Fig. 2D).

### Clinical sensitivity

We evaluated the clinical sensitivity of IgG_SP_ and IgM_SP_ assays in our RT-PCR-positive COVID-19 inpatient cohort where reliable information concerning the date and duration of symptom onset was available. Not surprisingly, we noted that the sensitivity of the assays increased proportionally with time following symptom onset, consistent with the developmental kinetics of a specific antibody response. The sensitivity of IgG_SP_ was 39% and 58% at 5 and 10 days following symptom onset (Table 1 and Fig. s1A). Extending the analysis of these patients between 16-20 days post symptom onset and beyond revealed a clinical sensitivity of IgG_SP_ assay was 96% and 98% respectively. As expected, clinical sensitivity of IgG_SP_ assay was much higher (74% within 10 days and 100% within 20 days) based on the RT-PCR confirmed date of diagnosis (Table s1; Fig. s1B).

**TABLE 1.** SARS-CoV-2 IgG_SP_ and IgM_SP_ positive agreement by days post symptom onset.

| **Days from symptom onset, DSO<sup>a</sup>** | **IgG<sub>SP</sub> positivity rate (%)** | **IgM<sub>SP</sub> positivity rate (%)** | **IgG<sub>SP</sub> or IgM<sub>SP</sub> positivity rate (%)** |
| --- | --- | --- | --- |
| <5 | 11/28 (39.3) | 11/28 (39.3) | 14/28 (50.0) |
| 6-10 | 46/83 (57.8) | 51/83 (55.4) | 55/83 (66.3) |
| 11-15 | 79/96 (82.3) | 76/96 (79.2) | 82/96 (85.4) |
| 16-20 | 42/44 (95.5) | 42/44 (95.5) | 44/44 (100) |
| >20 | 41/42 (97.6) | 40/42 (95.2) | 42/42 (100) |
| **Total** | 221/293 (75.4) | 220/293 (75.1) | 237/293 (80.1) |

<sup>a</sup> For RT-PCR confirmed SARS-CoV-2 cases.

Clinical sensitivity of IgM_SP_ test was comparable to that of the IgG_SP_ test both within 10 days and 20 days post symptom onset. At 20 days symptom onset and beyond, the sensitivity of IgM_SP_ test was 95% (Table 1), and 82% based on RT-PCR confirmed date of diagnosis (Table s1). Importantly, when IgG_SP_ and IgM_SP_ results were combined and analyzed for assay positivity in either format, the sensitivity reached 100% after 15 days from symptom onset and RT-PCR confirmed date (Table 1 and Table s1).

### Longitudinal assessment in COVID-19 inpatients

We next used our IgG_SP_ values over 360 specimens to assess the change in the units over every timepoint collected since the days post-infection confirmation by PCR (DPCR) with the data plotted as a frequency distribution mapping (Fig. s2A). Within the first 15 days of DPCR, the median values gradually increased and reached an average of 4466 AU/mL. A dramatic increase in the titer began to occur after 15 days (with a median average of 59396 AU/mL) (Fig. s2A). Beyond 27^th^ day, we noticed a decreasing trend in IgG_SP_ titers, however these results still remained above the positivity threshold (≥50 AU/mL). While a lower assay sensitivity was noted in the early period post symptom onset, it is notable that in some samples, the IgG_SP_ assay was able to detect positivity within the same timeframe as detectable SARS-CoV-2 RNA by RT-PCR. It is interesting to note that in three samples, IgG_SP_ assay was sensitive to detect the positivity in 0^th^ day, which is the date of infection confirmation by PCR.

In the same cohort, akin to IgG_SP_, we observed the overall tendency of IgM_SP_ to rise over the positivity threshold of 1.0 index value. Particularly, within the first week of symptoms, the distribution was highly inconsistent, and the maximum median index was only 4.345 (Fig. s2B). In contrast, we noted the IgM_SP_ distribution to progress regularly in tandem over the 2^nd^ week time with the median IgM_SP_ consistently doubling. This subsequently reached as high as ∼38 by the mid-2^nd^ week (17^th^ day). In 4 samples, IgM_SP_ was sensitive enough to detect the positivity in 0^th^ day of infection confirmation by PCR (Fig. s2B). While we were able to detect IgM_SP_ until 80^th^ day, the IgM_SP_ values were found to greatly oscillate beyond 23 days with a decreasing trend, however, still well above the cut-off 1.0 index value (Fig. s2B).

### IgG_NC_ assay cut-off for positivity much lower in exCOVID-19 (recovered) patients

This study included 16 blood samples from known COVID-19-recovered patients (recovered more than 3-11 months prior to sampling). Analysis of this group of patient samples with IgG assay specific for nucleocapsid protein (IgG_NC_), a main differentiator of immune responses to natural SARS-CoV-2 infection versus those to spike protein-based vaccines, showed index values ranging from 0.2 to <1.4 index value (Table 2; Fig. 3). This is significantly lower than manufacturer recommended cut-off index level of 1.4 used to detect active (early) infection. However, during waning humoral response (recovery period and post recovery), our data points out a value less than 1.4 and up to 0.2 index value may predict exCOVID-19 (recovered) (Table 2; Fig. 3). We therefore applied a cutoff below 0.2 for IgG_NC_ test to predict the true negatives (the absence of COVID-19), naïve group. This is important in the current setting of increasing numbers of COVID-19-recovered patients in the general population. More reliably, where a combination of tests is possible, the highest diagnostic accuracy of true negatives can be attained when combining a value of <0.2 index value for IgG_NC_ with <50 AU/mL for IgG_SP_ and <1.0 index value for IgM_SP_. 99% of the samples analyzed using this combination of results were confirmed as negative by RT-PCR or from patients who were in an early period of infection. By contrast, COVID-19 patients tested between 10-20 days of symptom onset and filtered with a combo of ≥1.4 index value for IgG_NC_ with≥ 50 AU/mL for IgG_SP_ and ≥1.0 index value for IgM_SP_ showed 99% positivity for PCR (Table 2). Subsequently we applied these filters wherever we analyzed naïve vaccinated and exCOVID-19 vaccinated groups.

**TABLE 2.** Threshold of different antibody assays used to predict the different conditions when used in combination in our dataset (filtered in).

| Conditions | IgG<sub>SP</sub> Assay | IgM<sub>SP</sub> Assay | IgG<sub>NC</sub> Assay |
| --- | --- | --- | --- |
| Infected (recent) | $\geq 50$ | $\geq 1.0$ | $\geq 1.4$ |
| Naïve | $< 50$ | $< 1.0$ | $< 0.2$ |
| Naïve Vaccinated | - | - | $< 0.2$ |
| Ex-COVID-19 | - | - | $0.2 - < 1.4$ |
| Ex-COVID Vaccinated | $\geq 50$ | $> 1.0$ | $0.2 - < 1.4$ |

**FIG 3.**
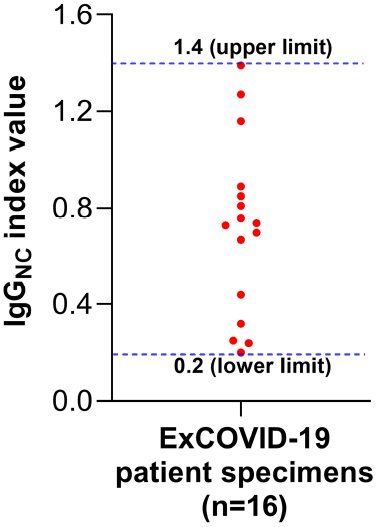
Derivation of new IgG_NC_ threshold based on the exCOVID-19 dataset. A total of 16 specimens from 11 unique COVID-19 recovered individuals were subjected for IgG_NC_ analysis against the vendor-recommended cut-off (≥ 1.4 index value).

### SARS-CoV-2 vaccination results in robust SARS-COV-2 spike protein-specific Ab responses

An exciting facet of this work is the inclusion of vaccinated patient samples in our analysis. Among those vaccinated, 91% received the Pfizer-BioNTech formulation (n=132) and 8% obtained the Moderna formulation (n=13). Ab responses in the vaccinated group (n=145) were compared to that of the unvaccinated group (n=424). The unvaccinated group included patients that were both RT-PCR positive and negative for SARS-CoV-2 infection. IgG_SP_ Ab response ranged from 0 to 49517 AU/mL, with a median of 6396 AU/mL (95% CI, 3814 to 9729) (Fig. 4A) in the vaccinated group. The median antibody level for vaccinated patients was significantly higher (∼6400-fold, p<0.0001) relative to those unvaccinated (Fig. 4A). Intriguingly, ∼18% (76/424) of unvaccinated patients had IgG_SP_ titers greater than positive detection threshold despite RT-PCR negativity. IgM_SP_ production was significantly (p<0.0001) higher in the vaccinated versus unvaccinated group, with a more modest median fold increase (∼23%, 1.34 vs 0.06) (Fig. 4B). Nearly 13% (53/424) of unvaccinated patients were above the IgM_SP_ positive threshold despite RT-PCR negativity.

**FIG 4.**
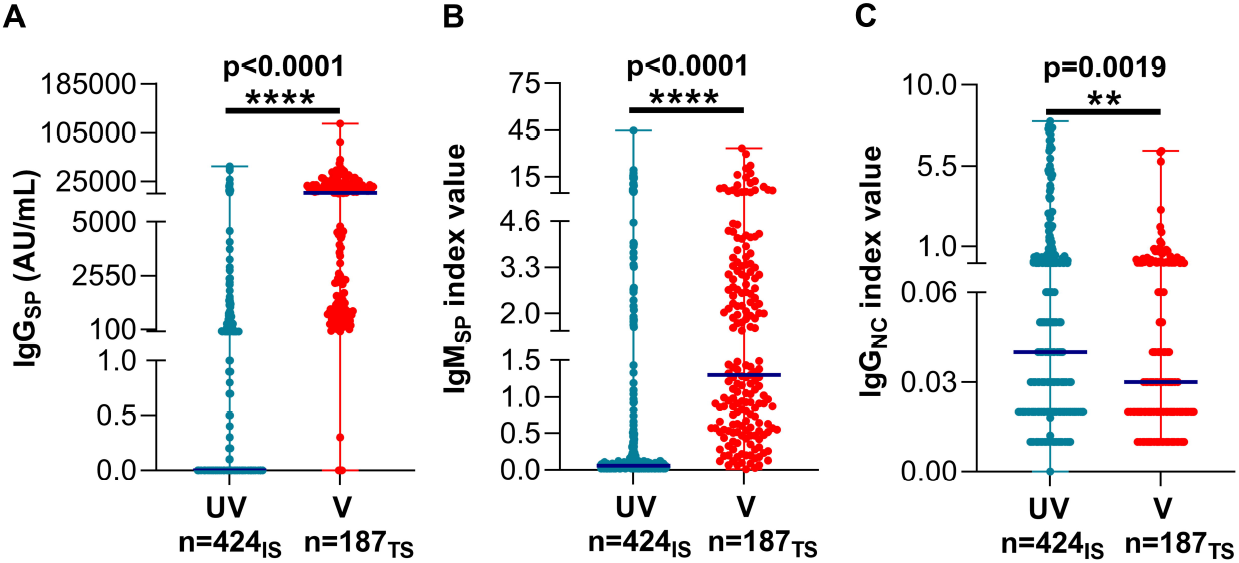
Humoral Ab response in vaccinated and unvaccinated cases. (A) Vaccination elicited-spike specific IgG response. (B) IgM_SP_ titers post-vaccination. (C) Nucleocapsid-specific IgG response in the unvaccinated and vaccinated. Cumulative dose 1 and dose 2 responses are shown. UV, Unvaccinated; V, Vaccinated; n, Number; IS, Individual samples; TS, Total specimens (that have two doses from same patients); Extended dark blue line indicates the median.

The identification of patients with positive IgG_SP_/IgM_SP_ results despite a lack of vaccination and historically negative RT-PCRs prompted us to investigate anti-nucleocapsid IgG titers in these patients as a marker of natural SARS-CoV-2 infection. Surprisingly, about 12% (51/424) were positive (≥1.4), leaving 26 (7%) patients with positive IgG_SP_. These patients could have silent infection (present or past) because they were also positive for either spike IgG or IgM. In a majority (82%; 347/424) of unvaccinated (UV) patients with no known SARS-CoV-2 exposure, the IgG_SP_ and IgM_SP_ titers were determined to be lower than the vendor-recommended cut-off.

Figure s3 illustrates the pattern of IgG_SP_ Ab titer relationships over the days following COVID-19 vaccination (DFD). This group (187 specimens from 145 unique individuals) included both RT-PCR verified cases, as well as subjects with no exposure to SARS-CoV-2. Within 10 days, the first dose yielded a minimal increase in vaccine-specific Ab response irrespective of formulation in 4 of 187 specimens. More than 54% (102/187) of the vaccinated group had 5-fold increase, 44% (81/187) had 10-fold increase, and 17% had 25-fold increase in the median IgG_SP_ titers when compared to the UV group (Fig. s3A). Samples up to 21 days are inclusive of the initial vaccine dose, and >21 days are inclusive of the booster. While an increase in titers following the 1^st^ dose of vaccination was clearly observed, not surprisingly its distribution was variable (Dark red rectangle; Fig. s3A). While the number of observations available at or less than 10 days following the 1^st^ dose was small, 4 of the individuals that were tested had titers under the positive threshold and 2 had titers over 45000 AU/mL. Clearly, after the 2^nd^ dose of vaccination, the IgG_SP_ titers was consistently high and comparatively exhibited a more uniform trend (Green Oval, Fig. s3A). Interestingly, 2 patients exhibited a null response following 21 days post vaccination. One of these patients was a HM (multiple myeloma) patient, while the medical history of the other is unknown.

In parallel, SARS-CoV-2-specific IgM_SP_ levels were also increased following vaccination in the naïve subjects (Fig. 4B & s3B). In particular, the distribution analysis demonstrated that following the primary dose of vaccination at week 1 and week 2, ∼71% of the subjects had values above the positive threshold (Fig. s3B). However, at 21 days and beyond post booster dose, IgM_SP_ levels again rose, and a noticeable over-the threshold cluster was seen compared to the primary vaccination (Fig. s3B). Although the number of Moderna-vaccinated recipients in this evaluation is modest, both the IgG_SP_ and IgM_SP_ responses appeared to be comparable to that of individuals who received the Pfizer-BioNTech formulation.

### Vaccination response in naïve population

Next, we compared the IgG_SP_ response after the first and booster dose in the naïve vaccinated group that was derived by filtering out for IgG_NC_ ≥ 0.2 based on Table 2 & Fig. 3. Following the first dose, we observed IgG_SP_ positivity with a median of 2217 AU/mL (95% CI, 0-44182), which was drastically increased by 8.2 fold following the booster to a median of 18272 AU/mL (at 98% CI, 11724-21750) (p<0.001) (Fig. 5A). The cumulative IgM_SP_ response after the primary dose of vaccination in the naïve subjects was found to be with a median of 1.1 index, which is above the positive cut-off (at 95% CI, 0.87-1.4) (Fig. 5B). This was further significantly increased (p=0.0236) by 1.7-fold to a median of 1.95 (at 98% CI, 1.16-3.3) following the booster dose (Fig. 5B). The IgG_NC_ remained unchanged between dose 1 and dose 2 in the naïve vaccinated group (Fig. 5C). This result suggests that there is a robust response after the booster dose in the naïve group.

**FIG 5.**
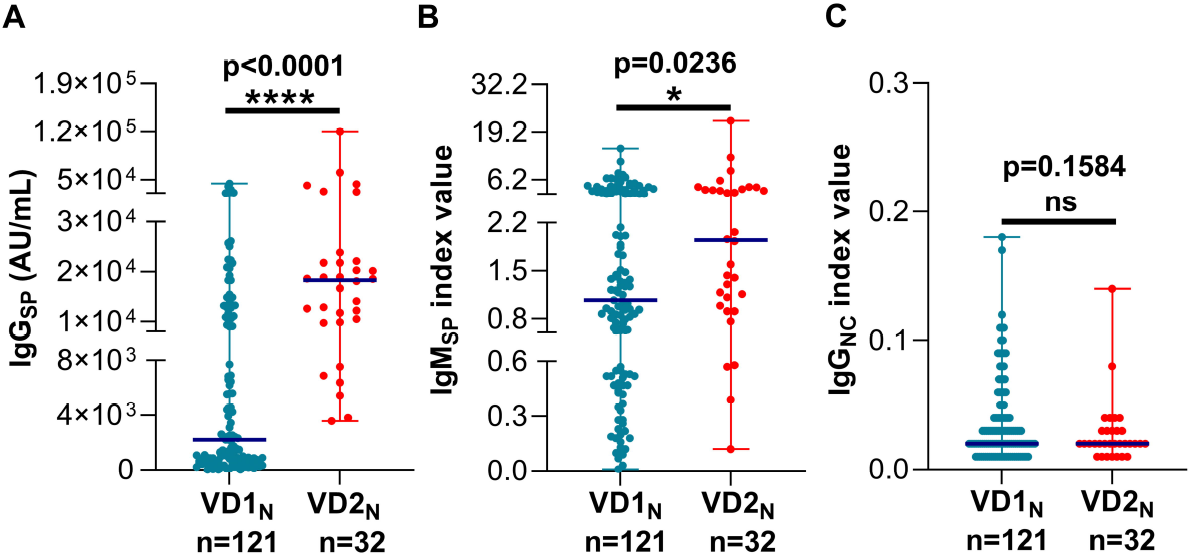
Comparison of Ab response post first and booster doses in naïve (non-COVID-19) subjects. (A) Evaluation of spike-specific Ab response in naïve (non-COVID-19) subjects following primary and booster dose of COVID-19 vaccine. (B) IgM_SP_ titers in naïve (non-COVID-19) subjects administered first and second dose of COVID-19 vaccine. (C) IgG_NC_ antibody levels following primary and booster vaccine dose in the naïve vaccinated subjects. V, Vaccinated; N, naïve (filtered in <0.2 for IgG_NC_), D1/D2, vaccine dose 1/2; n, Number; Extended dark blue line indicates the median.

### Vaccination response in exCOVID-19 (recovered) patients

Currently CDC recommends vaccination in those cohorts that are recovered from COVID-19 three months or prior. Therefore, understanding the immune response in this population is important. ExCOVID-19 group filter was derived from the known recovered patients (Table 2 & Fig. 3). The median IgG_SP_ titers following dose 1 and dose 2 in the exCOVID-19 group was found to be 17519 AU/mL and 20760 AU/mL, respectively (Fig. 6A). However, unlike the naïve vaccinated D1 vs D2 (Fig. 5A), the booster dose in the exCOVID-19 group displayed only a dampened response (Fig. 6A, lane 3 vs 4). A similar response to IgG_SP_ was noted for the IgM_SP_ in both the naïve vaccinated and exCOVID-19 vaccinated groups (Fig. 6B).

**FIG 6.**
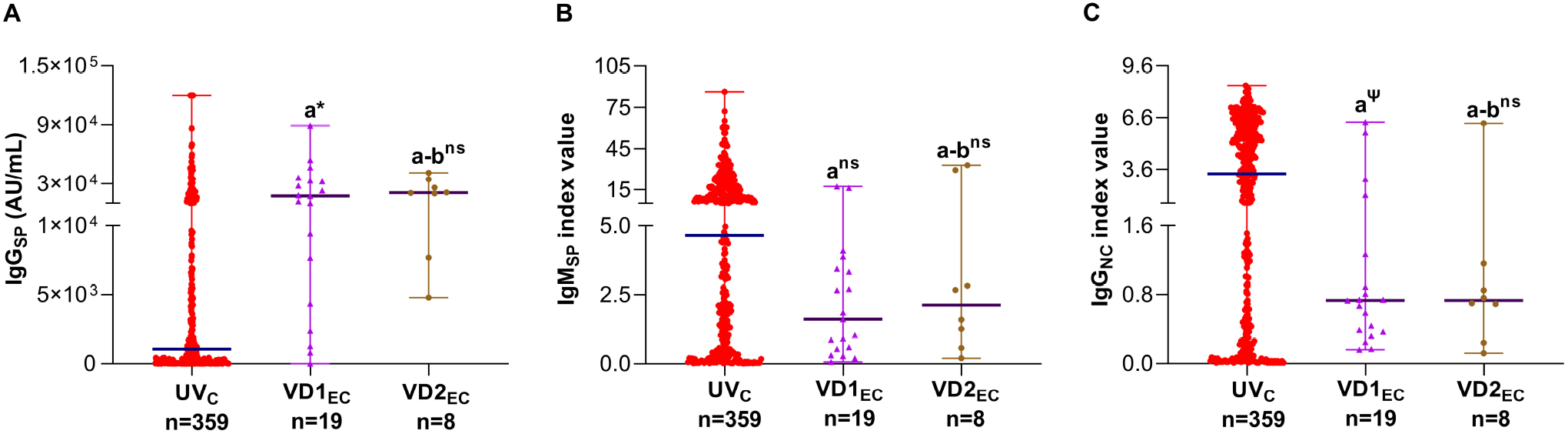
Comparison of Ab response following first and booster vaccine doses in exCOVID-19 (recovered) patients. (A) IgG_SP_ antibody titer in COVID-19 and exCOVID-19 (recovered) vaccinated subjects. (B) Assessment of IgM_SP_ response following first and second dose of vaccine in COVID-19 and exCOVID-19 (recovered) vaccinated subjects. (C) Comparison of IgG_NC_ antibody levels in COVID-19 and exCOVID-19 (recovered) vaccinated patients. UV, unvaccinated; V, vaccinated; C, COVID-19; EC, exCOVID-19 (recovered); D1/D2, vaccine dose 1/2; n, Number; Extended dark blue line indicates the median. a vs UV_C_; b vs VD1_EC_; *, p<0.0001; Ψ, p=0.0037; ns, not significant.

Next, we compared the vaccination response in exCOVID-19 against the active infection-based immune response (UV_C_). The infection response was confirmed by the IgG_NC_ levels that had a median of 3.3 (at 96% CI, 2.78 −4.02) that is 2.4-fold above the positive threshold cutoff (UV_C_; lane 1; Fig. 6C). Correspondingly, the median IgG_SP_ and IgM_SP_ titers were found to be 1046 (at 96% CI, 575.3 – 1518) and 4.65 (at 96% CI, 2.78 – 4.02), respectively (lane 1; Fig 6A & 6B). Both the doses robustly increased the IgG_SP_ response in relation to the infected group, while it was significant only after dose 1 (p<0.0001; lane 2 vs lane 1; Fig. 6A). In contrast, IgM_SP_ levels was significantly lower in the vaccinated group when compared to the infected group (2&3 vs 1, Fig. 6B). Interestingly, IgG_NC_ median titers remained much lower by 1-fold (0.73) than the positive cut-off of 1.4 suggesting IgG_NC_ response is infection-specific and not vaccination-specific. The vaccinated exCOVID-19 groups (D1 & D2) displayed significantly lower IgG_NC_ response when compared to the unvaccinated COVID-19 group and much below the positive threshold (lane 2&3 vs 1; Fig. 6C).

Within same patients, the antibody titers’ evaluation further confirmed the restrained response of booster dose in the exCOVID-19 recovered patients (lane 3 vs 4; Fig. s4A & s4B). Whereas the response was robust in the naïve vaccinated group (lane 1 vs 2; Fig. s4A & s4B). Within patient as well as within group comparison showed unchanged response for IgG_NC_ (Fig. s4C). As expected exCOVID-19 had higher nucleocapsid IgG levels when compared to the naïve (lane 3&4 vs lane 1&2, Fig. s4C). This result indicates a significant response for the first dose in the exCOVID subjects, while the booster dose appears to exert a restrained response.

## Discussion

SARS-CoV-2 serology has played very much a supporting role in the clinical diagnosis of COVID-19 compared to molecular detection of viral RNA. However, the recent availability of sensitive and specific antibody assays to quantitatively evaluate vaccination responses elevates the role of these important diagnostic tools at this juncture in the COVID-19 pandemic. In this work, we demonstrate that the Abbott Alinity quantitative IgG_SP_ and IgM_SP_ assays are sensitive and specific, and work well in a routine clinical setting. Additionally, these assays, when used in combination with the currently marketed IgG_NC_ assay, is able to measure and distinguish COVID-19 antibody responses from SARS-CoV-2 natural infection versus vaccination over a broad dynamic range.

The analytical performance of both the IgM_SP_ and IgG_SP_ assays met the acceptance criteria for the implementation of the assay in the clinical laboratory. The specificity of these assays was 100% in the samples from pre-COVID-19 period and there was no serological cross-reactivity in patients infected with other respiratory viruses (including seasonal coronaviruses). For IgM_SP_ assay, our results closely mirrored data provided by the manufacturer in the product insert. The high positive predictive value gained from minimal serological cross-reactivity in patients infected with other respiratory pathogens or autoimmune-associated antibodies is notable. Evaluation of the IgG_SP_ and IgM_SP_ assays utilizing RT-PCR positivity and days post symptom onset was to confirm the infection and the antibody response following the infection, respectively. The low level of sensitivity in samples collected between 5 and 15 days after symptom onset or PCR positivity is not surprising given the kinetics of antibody generation at these timepoints. However, the significant increase in sensitivity to greater than 95% beyond 15 days highlights the accuracy of these assays in the setting of an activated and productive humoral immune response. Therefore, these serologic assays, when used in conjunction with a molecular method which exhibits a lower degree of sensitivity at periods where serology is strongest, allows for a broad and thorough interrogation of a patient’s COVID-19 status.

Interestingly, when the IgG_SP_ with IgM_SP_ assays were analyzed in tandem, the sensitivity of the combination was improved than either the IgM_SP_ or IgG_SP_ assay individually with 100% sensitivity achieved after 15 days. Still, false-negatives in this paired approach were identified within 15 days, which could be due to host-specific factors, variations in immune responses, low antibody titers, or early SARS-CoV-2 infection.

While IgM is largely useful in determining the likelihood of a recent infection, the dynamics of how long this isotype remains detectable are ill-defined. In this study, we have detected IgM_SP_ for a protracted period (beyond 20 days and up to 80 days after symptom onset) with an increased sensitivity. This observation is more consistent with other studies that have detected IgM after ≥90 days following infection of SARS-CoV-2 (6,7). Interestingly, from our dataset, simultaneous detection of IgM and IgG was also noted, particularly, with the detection of IgG during early period, as well. This illustrates that seroconversion may occur simultaneously or shortly thereafter IgM_SP_ induction, in addition to the archetypal sequential class-switching responses (8).

The analysis of serum specimens from vaccinated patients is an exciting aspect of this work. These assays were able to detect and monitor vaccine-elicited antibody responses when compared to an unvaccinated patient group. The elevated levels of specific IgM_SP_ and IgG_SP_observed in this work is in agreement with the clinical trial data (9,10) for both currently available formulations of COVID-19 vaccines. Because it is not possible to parse if spike protein-specific antibodies arise because of vaccination or natural infection, we chose to utilize a currently marketed nucleocapsid-specific IgG test with the IgM_SP_/IgG_SP_ assays, which would only be positive in the setting of a natural infection.

Our results suggest changing the IgG_NC_ assay cut-off to ‘rule-in’ or ‘rule-out’ the ex-COVID-19. Specifically, our preliminary studies show a “grey zone” of IgG_NC_ index values (0.2 −1.4) in RT-PCR-confirmed COVID-19 recovered specimens (n=16 from 11 patients, recovery period within 3-11 months). While IgG_NC_ and IgG_SP_ assays demonstrated high clinical specificity and sensitivity independently using them in combination may allow distinguishing exCOVID-19 from vaccinated individuals in the field setting. Hence, we believe it is possible to expand the utility of the marketed IgG_NC_ assay to identify recovered individuals who are likely to present with waning antibody titers (11, 12). A similar revision of the IgG_NC_ threshold has been suggested to improve assay sensitivity for early stage (<6 days) of COVID-19 infection (13). Importantly, a model-based study recently proposed the use serostatus for prioritizing vaccination (14) and along these lines, the new cut-off range could improve the applicability with respect to identifying the previously infected and recovered. By identifying the relative abundance and characteristics of a population that is susceptible, infected, and recovered would be critical to better inform and devise the immunization roadmap. However, the cut-offs’ modification must be carefully considered in the clinical need and deployment setting. Studies to test the use of this cut-off algorithm on a larger scale are ongoing.

The combination of the new IgG_SP_ assay with the other assays, IgM_SP_ or IgG_NC_ can provide a rigorous evaluation of the vaccination and the antibody responses post-administration in the immunization setting. The deployment of the COVID-19 vaccine has faced many unique challenges due to its rapid development, hurried clinical trials and deployment. Limited knowledge concerning longitudinal outcomes, public uncertainty, and misinformation has created hesitancy among some of the public to receive this important immunization. These assays could aid in: (1) establishing the efficacy and kinetics of spike-specific antibody responses in infected or vaccinated populations, (2) help to identify infected individuals not previously identified by molecular testing, (3) evaluation of the effectiveness of the adjunct convalescent plasma therapy (CPT) that typically requires a quantitative assay-coupled neutralization assay. All of these mechanisms add to the breadth of knowledge concerning the COVID-19 vaccine and its effects, helping to fill in knowledge gaps and address the concerns of the public. While the US Centers for Disease Control and Prevention does not recommend serological testing to assess the COVID-19-driven immunity following mRNA vaccination, the use of a test that evaluates IgM/IgG to the nucleocapsid antigen for ascertaining prior infection in case of COVID-19 vaccination has been suggested (https://www.cdc.gov/vaccines/covid-19/info-by-product/clinical-considerations.html). Our work highlights an improved utility of a combination of antibody assays (spike and nucleocapsid) to accomplish this task.

Using these assays, while we observed an increased response to the vaccine following the primary dose (D1), the booster dose (D2) had a dramatic effect in the naïve subjects. Among vaccinated exCOVID-19 persons who had recovered from the infection, no significant difference is seen in IgG_SP_ and IgGM_SP_ titers following D2. However, D1 in exCOVID-19 recovered individuals elicited strong IgG_SP_ and IgG_SP_ responses of equivalent robustness to that of the D2 of the naïve group. D1 and D2 specimens from same individuals also confirmed this finding. The dramatic D1 response among exCOVID-19 recovered individuals could be ascribed to the natural SARS-CoV-2 infection functioning in a similar manner as acting as a priming dose. This observed response is supported by recent preprint reports that have demonstrated a single dose of mRNA vaccine in seropositive subjects is sufficient to elicit a robust immune response comparable to SARS-CoV-2 naïve subjects receiving two doses (15,16). These data raise important questions concerning whether a single dose is sufficient among previously SARS-CoV-2 infected and/or COVID-19 recovered individuals, however larger longitudinal studies are needed. Results of such work would have profound implications on vaccination strategies, particularly in the current setting of high COVID-19 vaccine demand with limited supply.

In sum, for the first time we demonstrate the clinical utility of the latest IgG_SP_ quantitative assay (under FDA approval) and IgM_SP_ test in detecting the vaccine response. In addition, the combination of IgG_NC_, IgG_SP_, and IgM_SP_ could be useful to distinguish prior infection status and vaccine related spike-protein humoral responses. Further, newly redefined IgG_NC_ threshold, when used in combination with the quantitative IgG_SP_ and IgMsp assays, can accurately identify COVID-19 recovered individuals allowing an improved diagnostic performance and enhancing public health surveillance and epidemiological initiatives.

## Data Availability

Yes

## Acknowledgements

We thank Abbott Diagnostics Division (IL, USA) for providing us SARS-CoV-2 specific Ab test kits for the validation of the assays in our facility and further clinical evaluations. We thank Drs. Chantale Lacelle, E Blair Solow and David Karp for providing their archived samples for specificity and cross-reactivity studies.

## Supplementary information

**TABLE s1** SARS-CoV-2 IgG_SP_ and IgM_SP_ positive agreement by days post RT-PCR verification.

**FIG.s1** Clinical sensitivity assessment. (A) Sensitivity of IgG_SP_ assay by days since PCR positivity (blue) and symptom onset (green). (B) Sensitivity of IgM_SP_ assay by days since PCR positivity (red) and symptom onset (green). PCR positive inpatient samples (n=360) that had symptom onset date were included in this analysis. DPCR, days post RT-PCR verification for PCR positivity; DSO, days from symptom onset.

**FIG.s2** Distribution of Ab response to SARS-CoV-2 shown for days post-PCR confirmation. (A) Frequency distribution mapping of IgG_SP_ results in COVID-19 PCR positive cases (n=355); (B) Frequency distribution analysis of IgM_SP_ results in COVID-19 PCR positive cases (n=355); DPCR, Date since PCR positivity; Broken red and blue lines, positive threshold.

**FIG.s3** Pattern of varied Ab response following vaccination. (A) IgG_SP_ results analyzed by frequency distribution (of cumulative dose 1 and dose 2 response) against days following vaccination; (B) Frequency distribution analysis (of cumulative dose 1 and dose 2 response) of IgM_SP_ results in vaccinated cases; DFD, Date following vaccination; Broken red and blue lines, positive threshold; Brown rectangle box, specified period post-dose 1 but prior to dose 2; Green circle, specific period after dose 2 vaccination.

**FIG.s4** Vaccine primary versus booster dose antibody response comparison within same subjects. (A) IgG_SP_ antibody titer following primary and booster dose of vaccination within same participant for naïve and COVID-19+ recovered groups. (B) Comparison of IgM_SP_ response within same subject in naïve and exCOVID-19 (recovered) group following primary and booster vaccine dose. (C) Same subject comparison for IgG_NC_ response in naïve and exCOVID-19 (recovered) group after primary and booster vaccine dose.

