## Supplemental Table 1 and Fig. s1, Fig. s2, Fig. s3, Fig. s4 for "Clinical evaluation of the Abbott Alinity SARS-CoV-2 spike-specific quantitative IgG and IgM assays in infected, recovered, and vaccinated groups"

**TABLE s1** SARS-CoV-2 $IgG_{SP}$ and $IgM_{SP}$ positive agreement by days post RT-PCR verification

| Days post RT-PCR verification, DPCR[a] | $IgG_{SP}$ positivity rate (%) | $IgM_{SP}$ positivity rate (%) | $IgG_{SP}$ or $IgM_{SP}$ positivity rate (%) |
|---|---|---|---|
| <5 | 67/118 (56.8) | 71/118 (60.2) | 79/118 (66.9) |
| 6-10 | 79/107 (73.8) | 79/107 (73.8) | 87/107 (81.3) |
| 11-15 | 55/67 (82.1) | 53/67 (79.1) | 58/67 (86.6) |
| 16-20 | 22/22 (100) | 22/22 (100) | 22/22 (100) |
| >20 | 44/45 (97.8) | 37/45 (82.2) | 45/45 (100) |
| **Total** | 270/359 (75.2) | 262/359 (72.9) | 291/359 (81.1) |

[a] For RT-PCR confirmed SARS-CoV-2 cases.

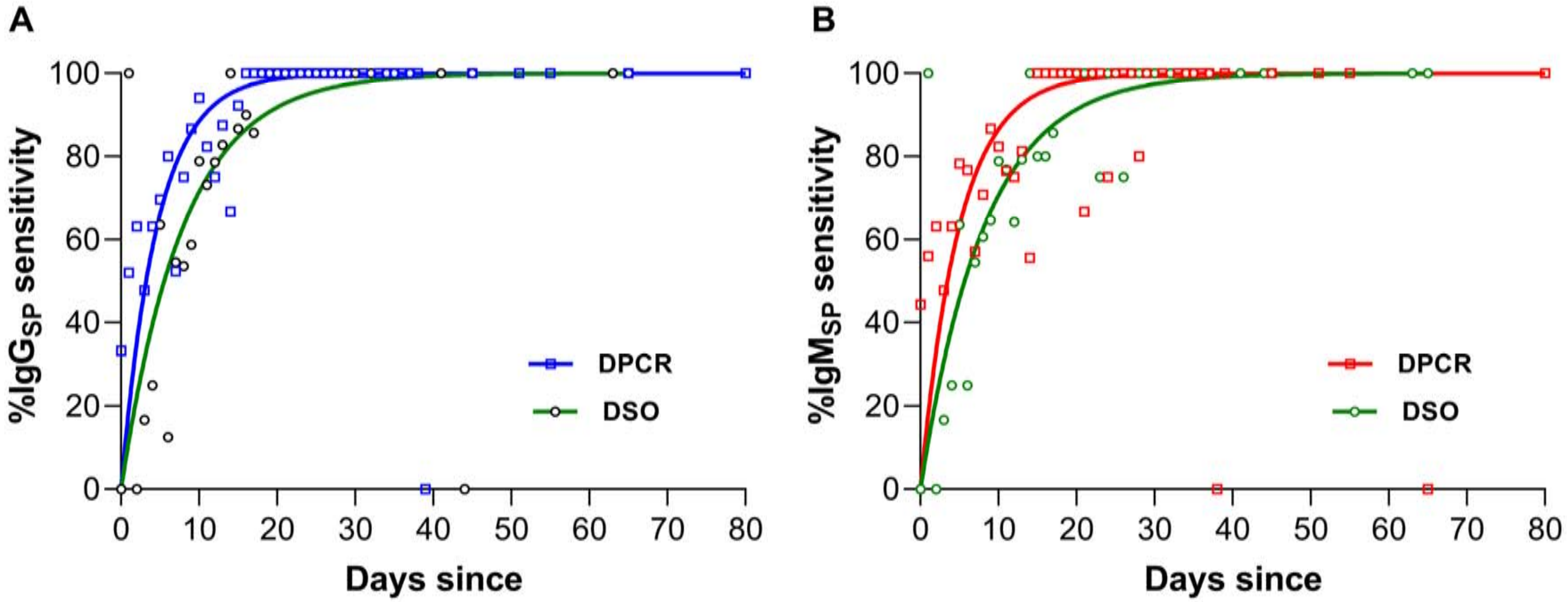
A
%IgG$_{SP}$ sensitivity
Days since
DPCR
DSO
B
%IgM$_{SP}$ sensitivity
Days since
DPCR
DSO

**Fig. s1**

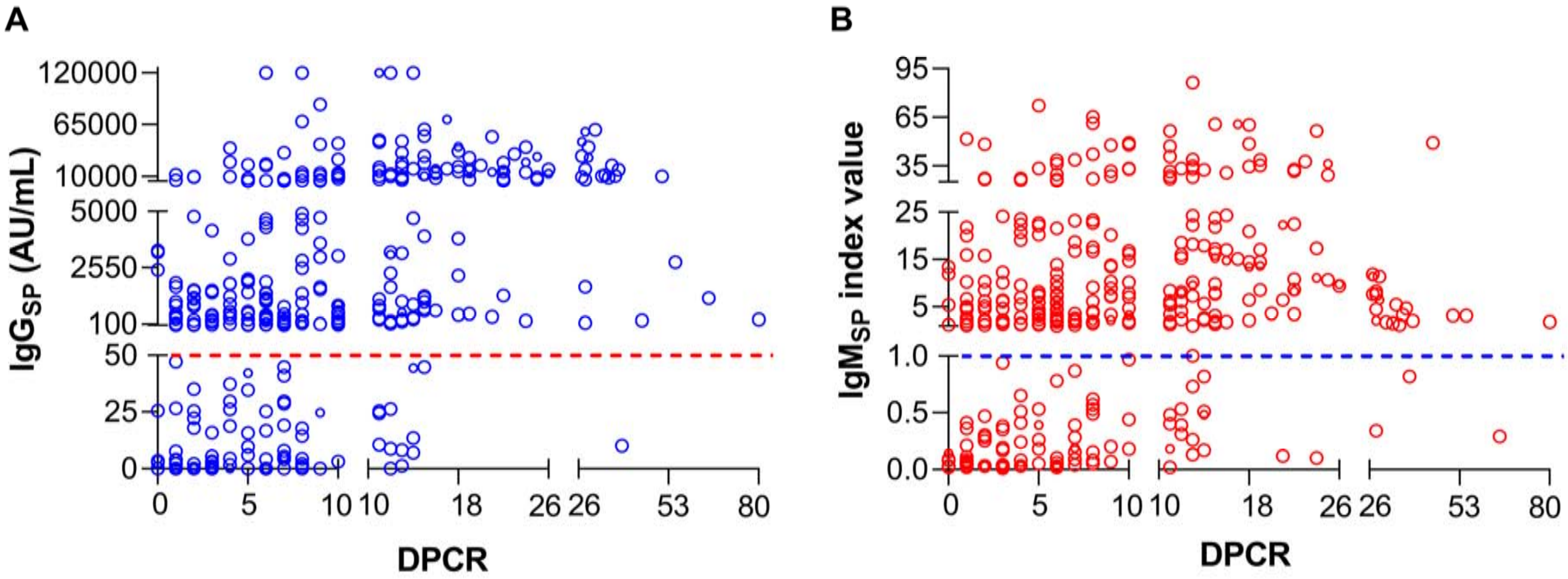

A
120000
65000
10000
5000
2550
100
50
25
0
IgG$_{SP}$ (AU/mL)
0
5
10
10
18
26
26
53
80
DPCR
B
95
65
35
25
15
5
1.0
0.5
0.0
IgM$_{SP}$ index value
0
5
10
10
18
26
26
53
80
DPCR


**Fig. s2**

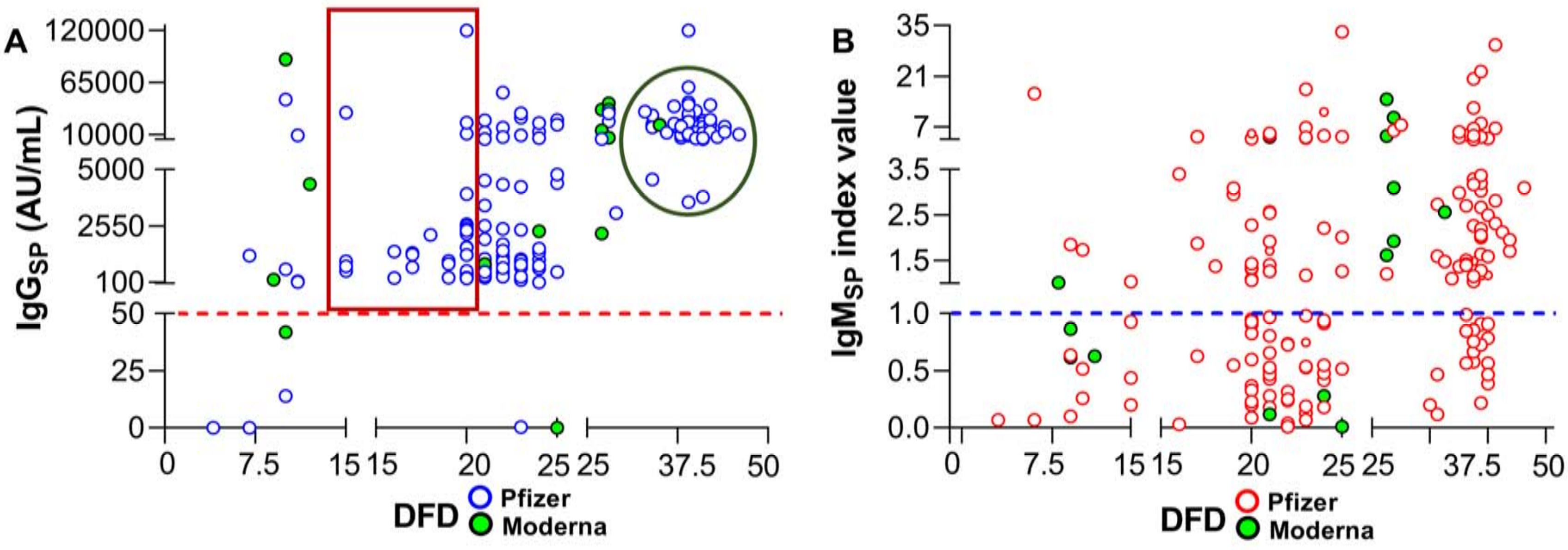

A
B
IgG$_{SP}$ (AU/mL)
IgM$_{SP}$ index value
120000
65000
10000
5000
2550
100
50
25
0
35
21
7
3.5
2.5
1.5
1.0
0.5
0.0
0
7.5
15
20
25
37.5
50
DFD
Pfizer
Moderna


**Fig. s3**

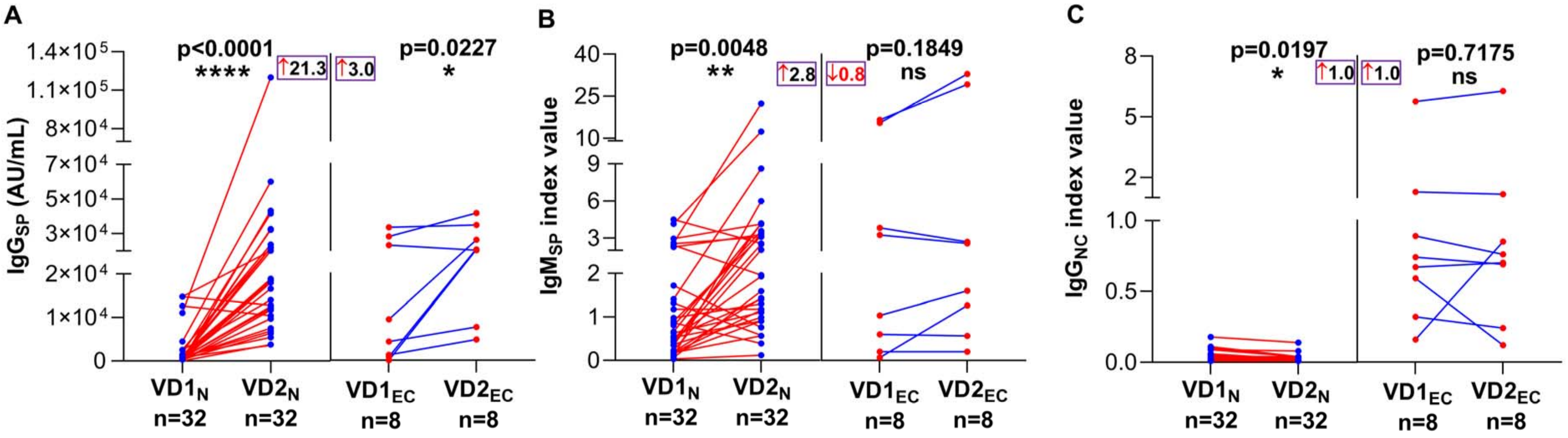
A
B
C
p<0.0001
****
p=0.0227
*
↑21.3
↑3.0
p=0.0048
**
p=0.1849
ns
↑2.8
↓0.8
p=0.0197
*
p=0.7175
ns
↑1.0
↑1.0
IgG$_{SP}$ (AU/mL)
IgM$_{SP}$ index value
IgG$_{NC}$ index value
VD1$_N$ n=32
VD2$_N$ n=32
VD1$_{EC}$ n=8
VD2$_{EC}$ n=8

Fig. s4
